# Growth charts for individualized evaluation of brain morphometry for preschool children

**DOI:** 10.1101/2021.04.08.21255068

**Authors:** Hongxi Zhang, Jia Li, Xiaoli Su, Yang Hu, Tianmei Liu, Shaoqing Ni, Haifeng Li, Xi-Nian Zuo, Junfen Fu, Ti-Fei Yuan, Zhi Yang

## Abstract

Brain development from 1 to 6 years-of-age anchors the rapid development of a wide range of functional capabilities. However, quantitative growth charts of typical development during this age period are lacking, preventing the identification of early brain abnormalities. Here we characterize the time-dependent individual differences of cortical thickness and subcortical volume in 340 typically developing children and construct regional growth curves for these brain morphological measures. The growth curves reflect four types of time-dependence for cortical thickness and subcortical volume metrics. At the individual level, the growth curve model provides percentiles for each brain region’s cortical thickness or volume during ages 1 to 6, allowing for individualized inferences of brain developmental status relative to the same-age population. The growth curves further demonstrate clinical utility potentials by identifying children with developmental speech and language disorders, achieving high accuracies on data collected on both 1.5T and 3T scanners. Our results fill the knowledge gap in brain morphometrics in a critical development period and provide an avenue for individualized brain developmental status evaluation, with demonstrated sensitivity and generalizability.

## Introduction

Brain development comprises complex morphological and volumetric changes [1,2]. Variations in the brain’s growth and regional differences emerge consistently during childhood and adolescence, contributing to inter-individual differences in general intelligence and functioning [3,4]. Such a life-span dynamic trajectory of both cortical and subcortical changes has been examined with a structural imaging approach [5,6]. Previous studies reported non-linear decreases in cortex gray matter volume and increases in white matter during the development period, mainly based on non-continuous or multiple site samples of children aged over eight or less than two years old [1,2,7–11]. This notable age gap reflects the challenge of recruiting healthy children between 1 to 6 years-of-age. Thus, the age-dependent morphometric dynamics of cortical and subcortical structures remain elucidated within this critical developmental period [12].

Two neuroimaging studies have covered this 1-6 age-span. One study reported cortical myelination changes reflected by myelin water fraction and T1 relaxation times [13]; the other study indicated that the white matter myelination reflected distinct neurodevelopmental processes from cortical thickness [14]. However, the annual change of cortical thickness and volume subcortical regions in this 1-6 age-span has not been quantified, and there is no model available to evaluate brain maturation.

To fill in this gap, we quantified typical year-by-year maturation in cortical thickness and subcortical volume based on brain imaging data from 340 children between 1 and 6 years old. We further developed growth curve models for all cortical and subcortical regions to characterize their age-dependence and to make inferences for individuals. We examined whether the models are useful for detecting abnormal brain development by training and testing classifiers to recognize children with language developmental disorders. The sensitivity and generalizability of the models were further examined using data from an independent scanner. The growth curve models reveal unique information that can help understand individual differences in the brain during this critical period and provide a novel tool for recognizing atypical structural brain development.

## Methods

### Participants

A total of 391 typical developing children (TDC) and 38 children with developmental speech/language disorder (DSLD) between one and six years old were recruited in the Department of Radiology, the Children’s Hospital affiliated to Zhejiang University School of Medicine, Hangzhou, China. The Medical Research Ethics committee at the Children’s Hospital approved the study. The parents of all participants signed the written informed consent form.

The inclusion criteria for TDC were: age between 1 and 6 years old; full-term (gestational age between 37 and 41 weeks) without any complicated perinatal course; referral for neuroimaging examination with indications including one episode of idiopathic febrile seizure, dizziness, headaches, facial or arm paralysis, trauma, short-term fever of unknown reason, and physical examination; typical results of neurological examination (examined by HL); free of current and past neurological or psychiatric disorders; and no evidence of genetic, metabolic, or infectious diseases. Exclusion criteria included: unable to complete the scans, poor image quality, and remarkable brain MRI radiological interpretation (examined by HX.Z).

The DSLD group was composed of patients referred to a neuroimaging examination with an indication of developmental speech/language disorder. The clinical diagnosis of DSLD was confirmed by a pediatric neurologist (H.L.), following criteria for language development delays[15,16]. The inclusion criteria were: age between 1.5 and 6 years old; full-term (gestational age between 37 and 41 weeks) without any complicated perinatal course; unable to produce a single word before 18 months of age, or produced less than 30 words after age 24 months, or expressing fewer than 3/5 linguistic structures for boys/girls after 30 months, or unable to express two-word phrases after 36 months of age; and no evident brain structural abnormality on conventional MRI. Participants with any of the following were excluded: autism spectrum disorder, intellectual disability, hearing deficit, phonological production deficit, or other sensory deficits; tics disorders, coprolalia syndrome, attention-deficit/hyperactivity disorder, specific learning disability, anxiety disorders, depressive disorders, or seizures. A senior radiologist (HX.Z) further assessed clinical scans (both T1 and T2 weighted images) to exclude remarkable brain structure abnormalities and myelination abnormalities.

### MRI acquisition

Twenty-six DSLD participants and 303 TDC were scanned with a Siemens 1.5T MRI scanner (Magnetom Avanto, Siemens Healthcare, Erlangen, Germany). Besides regular T1- and T2-weighted clinical scans, high-resolution T1-weighted 3D images were acquired using a magnetization prepared rapid gradient echo (MPRAGE) sequence with the following parameters: TR = 1910 ms, TE = 3.06 ms, TI = 1100ms, flip angle = 15°, FOV = 192 mm × 192 mm, data matrix = 256 × 256, spatial resolution = 0.8 × 0.8 × 1.0 mm^3^. The participants who could not cooperate with examinations due to their young age were sedated with 10% chloral hydrate (50mg/ml) orally or by enema before scanning.

The other 66 TDC and 13 DSLD participants were scanned with a Philips Achieva 3T TX MRI scanner (Philips Healthcare, Best, The Netherlands). Besides regular T1- and T2-weighted clinical scans, high-resolution T1-weighted 3D images were acquired using a 3D Turbo Field Echo (3D-TFE) sequence with the following parameters: TR = 2000 ms, TE = 3.7 ms, TI = 800ms, flip angle = 8°, FOV = 256 mm × 256 mm, data matrix = 320 × 320, 180 slices, spatial resolution = 0.8 × 0.8 × 2.0 mm^3^.

### Image processing

The clinical scans were first interpreted to exclude participants who exhibited remarkable brain structural abnormality. The T1-weighted 3D images of the remained participants were visually checked for head motion artifacts. Images with obvious artifacts were further dropped from further analyses.

A total of 292 TDC completed MRI scans on the 1.5T scanner, among which 11 were excluded due to remarkable brain abnormalities, and 16 were excluded due to poor image quality based on visual inspections 265 were included in further analyses. The sample sizes from age 1 to age 6 were: 52 (22 females), 49 (20 females), 47 (19 females), 34 (15 females), 43 (18 females), and 40 (24 females), respectively. Supplementary Table 1 presents a summary of the indications when referred for neuroimaging examination for the 265 TDC. The most common indications were idiopathic febrile seizure, headache, and physical examination. These three types of indications accounted for 80.4% of the sample.

A total of 26 developmental speech and language disorder (DSLD) patients, who received “no obvious abnormal” radiology diagnoses, completed brain scans on the 1.5T MRI scanner, among which six were excluded due to low image quality, remaining 20 DSLD for further analyses. There were six females and 14 males. The mean age was 3.03, with a standard deviation of 1.29 years. To form a control group for the 20 DSLD patients, we selected 20 people from the 265 TDC matched with the DSLD patients one-on-one by sex and age. The mean age of this control group was 2.97, with a standard deviation of 1.37 years.

Sixty-six TDC and 13 DSLD participants completed MRI scans on the 3T scanner, but 11 TDC and 3 DSLD participants were excluded due to poor image quality or remarkable brain abnormalities, remaining 55 TDC and 10 DSLD for further analyses. The sample sizes of TDC from age 1 to age 6 were: 16 (9 females), 11 (7 females), 10 (7 females), 3 (2 females), 5 (2 females), and 10 (7 females). Supplementary Table 2 presents a summary of the indications in the referral for neuroimaging examination for the 55 TDC included in the analyses. The most common indications were idiopathic febrile seizure, headache, and physical examination, which accounted for 87.3% of the sample.

Supplementary Figure 1 presents a flowchart of the following imaging analysis steps. Brain extraction results from CAT12, volbrain [17], ANTs [18], and FreeSurfer [19] were compared. Based on a visual check of all images, the brain masks generated by the volbrain were chosen as the brain extraction masks. Next, the TDC-1.5T group was split into a training set containing 245 participants and a test set of 20 participants that best matched the age and sex of the 20 DSLD participants.

We divided the TDC in the training dataset into six non-overlapping age groups (i.e., 1-2, 2-3, 3-4, 4-5, 5-6, and 6-7 years old). A brain image template was constructed for each group using ten male and ten female participants who were randomly selected, following ANTs’ multivariate template construction procedure [20]. This procedure has been used in previous studies to generate population-specific brain templates [21,22]. Then the multivariate joint label fusion procedure in ANTs [23], combined with a set of 15 manually labeled brains, was applied to each age-specific template image to generate six image segmentation prior probability images, representing gray matter, white matter, cerebrospinal fluid (CSF), subcortical structures, brain stem, and cerebellum, respectively. Supplementary Figure 2 presents the resultant age-specific brain templates and corresponding prior probability images.

With these age-specific prior brain tissue probability images and brain templates, the n-tissue segmentation tool in ANTs was applied to obtain white matter masks for all participants. The white matter segmentation quality for all participants was visually checked, and all images yielded acceptable white matter masks. The cortical thickness and subcortical volume statistics for each participant were estimated using FreeSurfer. Specifically, the denoised brain structure images from ANTs’ N4 denoising step were input to the FreeSurfer “recon-all” pipeline. The corresponding white matter and brain extraction masks obtained previously were injected into the pipeline to improve its performance in constructing surface models for this very young sample. The FreeSurfer “recon-all” pipeline parcellated the cerebral cortex into 62 (31 per hemisphere) anatomical regions defined by the Desikan-Killiany Atlas and parcellated subcortical structures into 20 regions (excluding ventricles and brain stem) per the default ‘aseg’ atlas in FreeSurfer. The “recon-all” pipeline calculated regional mean cortical thickness, subcortical region volume, and volume of a variety of brain tissues such as cerebral white matter, subcortical gray matter, cerebellar white matter, cerebellar cortex. These metrics were used in further analyses.

### Age-group comparisons

In the training dataset (n = 245), we first applied a linear model to each of the above brain morphometrics to examine the effects of age, sex, and their interactions. To avoid the potential impact of outliers, we cleaned the data by setting the values outside the [Q_0.25_ – 1.5 IQR, Q_0.75_ + 1.5IQR] range to the closest boundaries of this range, where Q_0.25_ and Q_0.75_ are the first and third quartiles, and IQR indicates the distance between Q_0.25_ and Q_0.75_. We then characterized the age-dependence of the metrics by 1) calculating the change rate for each group from age 2 to age 6, relative to age 1, and 2) examining year-by-year changes using independent sample t-tests. The significance was corrected for multiple comparisons using the false-discovery rate (FDR) approach.

### Growth curve model

We constructed a growth curve model for each of the metrics to model the age-dependence of both typical value and individual variability of the brain morphometrics. Furthermore, the models can evaluate relative positions of a given individual among the same-age population, analogous to the developmental score for height or weight of a child. The model construction procedures were largely consistent with those used for constructing height/weight growth curves by the World Health Organization [24]. Technical details, as well as differences from the WHO procedures, are described below. The following procedure was applied to each metric. For those exhibiting significant sex differences, additional sex-specific growth curve models were constructed.

#### GAMLSS model

The Generalized Additive Model for Location, Scale, and Shape (GAMLSS) was adopted to fit the growth curves. This model is a distributional regression approach, allowing for modeling age-dependent changes for all probability distribution parameters. Mathematically, the model can be expressed as:

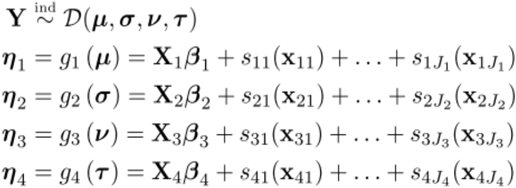

In this formulation, X_i_ and x_ij_ are subsets of the exploratory variable, i.e., TDC participants’ precise age. Y is the response variable, i.e., the value of a given regional metric. ***D*** is a probability distribution, and µ, *σ, ν*, and *τ* are possible parameters. The number and meaning of these parameters vary with the specific form of the distribution ***D*** determined in the model fitting step. *g*1 to *g*4 are link functions to map the parameters onto the entire real axis, and s_ij_ are nonparametric smoothing functions of explanatory variables. X_i_*β*_i_ and s_ij_(x_ij_) are additive terms. Model fitting was implemented using the R package “GAMLSS”.

#### Choice of distribution

To choose a proper distribution (***D***) for the current dataset, four distributions were examined, including Box-Cox power exponential, Box-Cox t, Box-Cox normal, and Johnson’s SU. The training samples were divided into six non-overlapping age groups (1-2, 2-3, 3-4, 4-5, 5-6, and 6-7 years old), and model fitting was conducted within each age group without considering additive terms. Per the Akaike Information Criterion (AIC), the Box-Cox normal distribution exhibited the best goodness of fit for over 60% of all six age groups’ metrics. We, therefore, adopted this distribution for model fitting.

#### Choice of additive terms

Two types of additive terms were compared: second-order polynomial and second-order fractional polynomial. The polynomial term can be expressed as: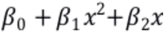, and the fractional polynomial term can be expressed as: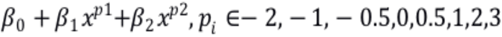. The AIC was compared when using these terms to fit the three parameters of Box-Cox normal distribution with three parameters, µ, *σ*, and *ν*, representing mean, standard deviation, and skewness. The second-order polynomial term yielded smaller AIC for all regional metrics, and thus this additive term was adopted in the GAMLSS model.

#### Model fitting

The following model fitting procedure was applied:

1. Construct model 1 to fit only µ (see the GAMLSS formula above).
2. Conduct Q-test on *σ, ν* in model 1. Q-test examines the normality of the residuals within a range of an independent variable and has been used to construct WHO Child Growth Standards to detect parameter misfit. If any parameter’s Q-test p-value is less than 0.05, model 2 will be constructed to fit µ and *σ*.
3. Conduct Q-test on *ν* in model 2 if it is constructed. If *ν*’s Q-test p-value is less than 0.05, model 3 will be constructed to fit µ, σ, and *ν*.
4. Compute generalized AIC (GAIC) for the available models. GAIC=-2L+km, where L is the likelihood, m is the number of parameters, and k was set to 5, given the relatively limited sample size. The model with the smallest GAIC was chosen as the final model.

### Clustering analyses

We conducted hierarchical clustering analyses to reveal the representative development patterns of the cerebral regions’ thickness and the volume of the subcortical regions. Since 81 unique values of precise age uniformly distributed between 1 to 7, we extracted the fitted median values for the 81 precise ages and transformed them into Z scores. Euclidean distance between the regions was then computed based on the 81-dimensional vectors. The hybrid hierarchical clustering algorithm, implemented in the R package “hybridHclust” [25], was then applied to this distance matrix. Briefly, this method includes three steps:

1. Use bottom-up clustering to find the mutual clusters, each of which is a group of sufficiently close points to each other and distant from all other points.
2. Perform a constrained top-down clustering that retains the mutual clusters.
3. Perform a top-down clustering within each mutual cluster.

We chose the numbers of clusters based on the “knee points” on the inter-cluster distance vs. the number of clusters plots to visualize the representative developmental patterns. This procedure was performed for the cortical thickness, and subcortical volume results separately. The dendrograms were cut into clusters according to the chosen number of clusters. The within-cluster growth curves were normalized to relative changes against age 1 and averaged to represent the clusters’ typical patterns.

### Recognizing DSLD using brain morphometrics

To evaluate the utility of the growth curve models, we examined whether TDC and DSLD participants could be accurately classified using the development scores (i.e., the percentiles among the same-age population) derived from the growth curve models. For those metrics showing significant age-differences, the development scores were derived from the corresponding sex-specific growth curves. Four classifiers, based on regularized discriminant analysis (RDA), as implemented in the R package “klaR” [26], were trained to recognize DSLD from TDC participants. The four classifiers included the regional cortical thickness, subcortical volume, volume of brain tissues, and all metrics as features, respectively. The performance of the classifiers was evaluated using leave-one-out cross-validation. Classification accuracy and the area under the receiver operator curves (AUC) were calculated to indicate the classifiers’ performance.

To further examine the sensitivity and generalizability of the growth curve models, we applied the growth curve models constructed using the data from the 1.5T scanner (n =245) to the images from the 3T scanner to derive a percentile value among the same-age population for each region in each participant. These data were then inputted to the classifiers trained using the 20 TDC and 20 DSLD participants scanned on the 1.5T scanner. This procedure formed an independent test of the performance of the classifier. We calculated the AUC of the classifiers to indicate their performance.

### Data availability

We shared the brain templates for children from 1 to 6 years old (3D nifty files), the growth curve models of all brain regions, and the code to perform the analysis in a public open-science repository: (https://osf.io/fm7cq/?view_only=9716e89f09e04b4bb2b4f0323ab2b684). The original and processed imaging data are available on reasonable request.

## Results

### Age-difference of brain morphometrics

#### Brain tissue volumes

Using a linear model, we examined the main effects of sex and age and their interactions on the volume of a series of brain tissues, including brain volume (excluding ventricles), total gray matter volume, total cortex volume, total cerebral white matter volume, subcortical gray matter volume, cerebellar white matter volume, and cerebellar cortex volume. There was no metric showing significant sex by age interaction effects. After false-discovery rate (FDR) correction (q<0.05), the brain volume, total cortex volumes, total cerebral white matter volumes, and total gray matter volume exhibited significant differences between male and female participants.

Table 1 displays the age-group differences and the change rate of the volume of brain tissues. According to these results, ages 2-3 exhibited the most significant increase in all types of brain tissues. Age 4 also showed a significant volume increase in cerebral white matter, cortex, total gray matter, and whole-brain (excluding ventricles). Supplementary Table 3 presents age-group differences and change rates for male and female children separately for those brain tissues showing significant sex-difference. The observations in the pooled samples remained, while after multiple-comparisons correction, only male children exhibited a significant increase in the volume of the brain (excluding ventricles), total gray matter, total cortex, and total cerebral white matter at age 4.

**Table 1.**
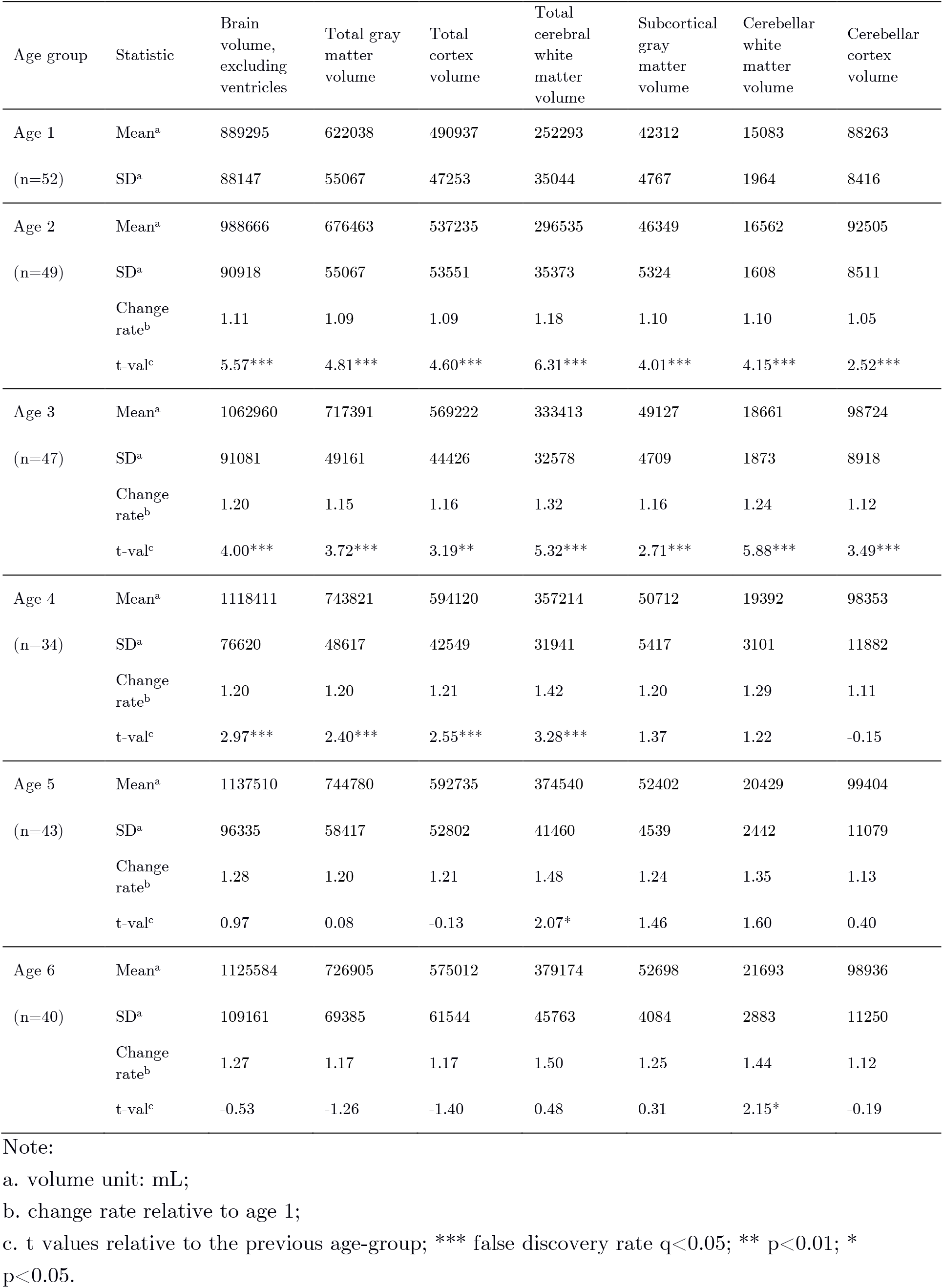
Age-dependence of brain tissue volumes.

| Age group | Statistic | Brain volume, excluding ventricles | Total gray matter volume | Total cortex volume | Total cerebral white matter volume | Subcortical gray matter volume | Cerebellar white matter volume | Cerebellar cortex volume |
| --- | --- | --- | --- | --- | --- | --- | --- | --- |
| Age 1 | Mean<sup>a</sup> | 889295 | 622038 | 490937 | 252293 | 42312 | 15083 | 88263 |
| (n=52) | SD<sup>a</sup> | 88147 | 55067 | 47253 | 35044 | 4767 | 1964 | 8416 |
| Age 2 | Mean<sup>a</sup> | 988666 | 676463 | 537235 | 296535 | 46349 | 16562 | 92505 |
| (n=49) | SD<sup>a</sup> | 90918 | 55067 | 53551 | 35373 | 5324 | 1608 | 8511 |
|  | Change rate<sup>b</sup> | 1.11 | 1.09 | 1.09 | 1.18 | 1.10 | 1.10 | 1.05 |
|  | t-val<sup>c</sup> | 5.57*** | 4.81*** | 4.60*** | 6.31*** | 4.01*** | 4.15*** | 2.52*** |
| Age 3 | Mean<sup>a</sup> | 1062960 | 717391 | 569222 | 333413 | 49127 | 18661 | 98724 |
| (n=47) | SD<sup>a</sup> | 91081 | 49161 | 44426 | 32578 | 4709 | 1873 | 8918 |
|  | Change rate<sup>b</sup> | 1.20 | 1.15 | 1.16 | 1.32 | 1.16 | 1.24 | 1.12 |
|  | t-val<sup>c</sup> | 4.00*** | 3.72*** | 3.19** | 5.32*** | 2.71*** | 5.88*** | 3.49*** |
| Age 4 | Mean<sup>a</sup> | 1118411 | 743821 | 594120 | 357214 | 50712 | 19392 | 98353 |
| (n=34) | SD<sup>a</sup> | 76620 | 48617 | 42549 | 31941 | 5417 | 3101 | 11882 |
|  | Change rate<sup>b</sup> | 1.20 | 1.20 | 1.21 | 1.42 | 1.20 | 1.29 | 1.11 |
|  | t-val<sup>c</sup> | 2.97*** | 2.40*** | 2.55*** | 3.28*** | 1.37 | 1.22 | -0.15 |
| Age 5 | Mean<sup>a</sup> | 1137510 | 744780 | 592735 | 374540 | 52402 | 20429 | 99404 |
| (n=43) | SD<sup>a</sup> | 96335 | 58417 | 52802 | 41460 | 4539 | 2442 | 11079 |
|  | Change rate<sup>b</sup> | 1.28 | 1.20 | 1.21 | 1.48 | 1.24 | 1.35 | 1.13 |
|  | t-val<sup>c</sup> | 0.97 | 0.08 | -0.13 | 2.07* | 1.46 | 1.60 | 0.40 |
| Age 6 | Mean<sup>a</sup> | 1125584 | 726905 | 575012 | 379174 | 52698 | 21693 | 98936 |
| (n=40) | SD<sup>a</sup> | 109161 | 69385 | 61544 | 45763 | 4084 | 2883 | 11250 |
|  | Change rate<sup>b</sup> | 1.27 | 1.17 | 1.17 | 1.50 | 1.25 | 1.44 | 1.12 |
|  | t-val<sup>c</sup> | -0.53 | -1.26 | -1.40 | 0.48 | 0.31 | 2.15* | -0.19 |

Note:
a. volume unit: mL;
b. change rate relative to age 1;
c. t values relative to the previous age-group; \*\*\* false discovery rate $q < 0.05$ ; \*\* $p < 0.01$ ; \* $p < 0.05$ .

#### Cortical thickness

There was no region showing a significant interaction effect between age and sex or the main effect of sex on cortical thickness (after FDR correction). Figure 1 shows maps of cortical thickness across 1-6. For most cortical regions, the cortical thickness was between 2 and 4 mm and showed changes across ages. To quantify the age-dependence of cortical thickness, we examined the age-group difference of mean thickness of every cortical region. Figure 2A shows maps of mean annual change rates across 2-6 years old, relative to the first year. For most regions, the cortical thickness exhibited a monotonous downward trend. At age 6, the cortical thickness of the bilateral orbitofrontal cortex, bilateral cuneus, and bilateral pericalcarine gyrus was thinner than 90% of age 1. For the precentral gyrus, entorhinal cortex, and parahippocampal gyrus, the cortical thickness first increases and decreases in age 4 and age 5. Still, the cortex in these regions was not thinner than the first year for all age groups. The cortical thickness changes reflected symmetry between the two hemispheres, i.e., the corresponding regions between hemispheres showed similar changing trends. Figure 2C shows t maps of the year-by-year difference of all cortical regions. The white dots mark a significant year-by-year difference (multiple-comparisons corrected using FDR, q < 0.05). These results showed that most cortical regions’ significant changes appeared in age 2, age 3, and age 6.

**Figure 1.**
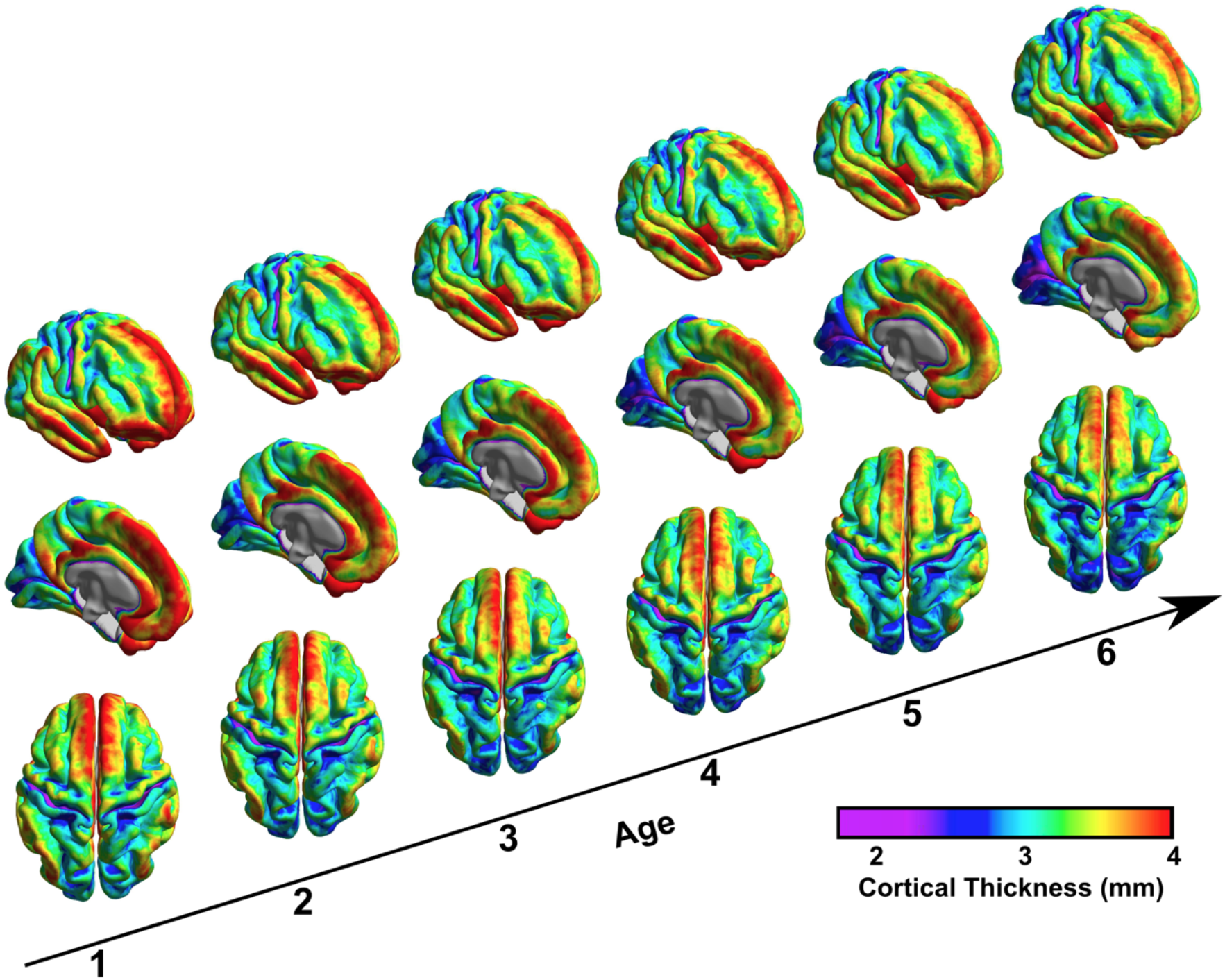
Dynamic maps of cortical thickness. The colors indicate cortical thickness. Most cortical regions become thinner from 1 to 6 years old. The medial frontal cortex, anterior cingulate cortex, and the parietal cortex exhibit fast changes during this period.

**Figure 2.**
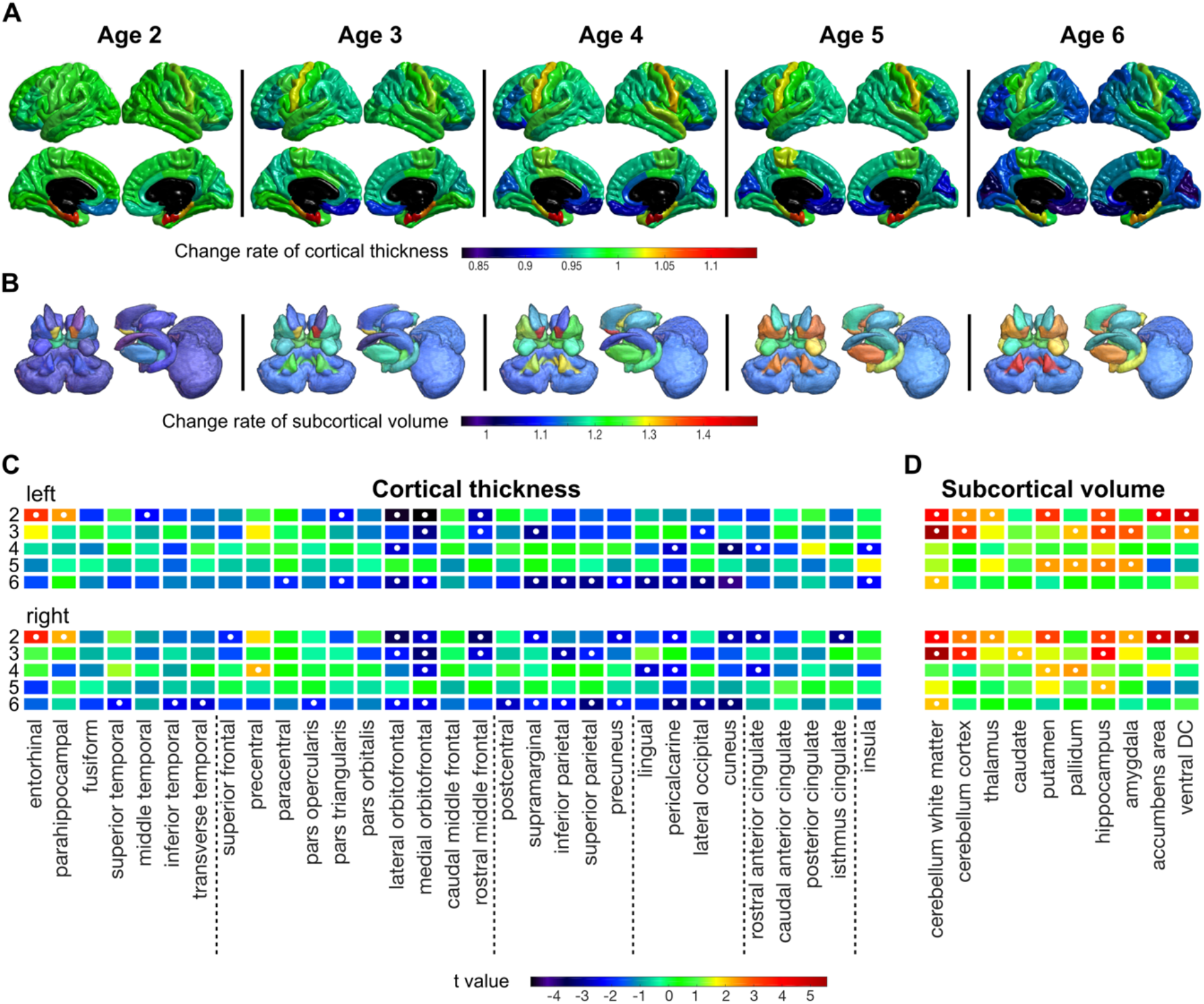
Annual changes in cortical and subcortical regions. A. Change rate of cortical thickness, relative to age 1. While most regions become thinner, the cortical thickness of the precentral gyrus, entorhinal cortex, and parahippocampal gyrus increases and decreases. B. Change rate of volume of subcortical regions, relative to age 1. Most subcortical regions’ volume increases with age, and some regions are 30% larger than the first year. C. Year-by-year difference of cortical thickness. The colors represent t-values comparing the current year with the year before. The white dots indicate significant differences after multiple-comparison correction using a false-discovery rate < 0.05. D. Year-by-year difference in the volume of subcortical regions. The colors represent t-values comparing the current year with the year before. The white dots indicate significant differences after multiple-comparison correction using a false-discovery rate < 0.05.

#### Subcortical volume

There was no significant interaction effect between age and sex on subcortical regions’ volume after FDR correction. The volume of bilateral thalamus exhibited significant sex difference after FDR correction: left thalamus: (F = 15.03, p = 0.00014), right thalamus (F=8.47, p = 0.004). Table 2 presents the mean and standard deviation of the volume of subcortical regions for each age group.

**Table 2.** The volume of subcortical regions between 1 and 6 years old.

|  |  | Age 1 |  | Age 2 |  | Age 3 |  | Age 4 |  | Age 5 |  | Age 6 |  |
| --- | --- | --- | --- | --- | --- | --- | --- | --- | --- | --- | --- | --- | --- |
|  |  | Mean | SD | Mean | SD | Mean | SD | Mean | SD | Mean | SD | Mean | SD |
| Cerebellum white matter | L | 7362 | 878 | 8055 | 839 | 9079 | 938 | 9393 | 1618 | 9927 | 1131 | 10511 | 1417 |
|  | R | 7721 | 1129 | 8507 | 879 | 9582 | 976 | 9999 | 1586 | 10460 | 1263 | 11032 | 1281 |
| Cerebellum cortex | L | 43842 | 4160 | 46010 | 4301 | 49157 | 4308 | 48688 | 5143 | 49159 | 5346 | 48935 | 5705 |
|  | R | 44422 | 4316 | 46494 | 4292 | 49460 | 4425 | 49768 | 4902 | 50132 | 5588 | 50000 | 5634 |
| Thalamus | L | 5720 | 723 | 6108 | 924 | 6371 | 711 | 6561 | 763 | 6548 | 672 | 6590 | 538 |
|  | R | 5895 | 788 | 6292 | 1002 | 6588 | 748 | 6550 | 668 | 6816 | 739 | 6824 | 701 |
| Thalamus (Male) | L | 6215 | 808 | 6470 | 919 | 6640 | 692 | 6744 | 495 | 7010 | 709 | 6905 | 716 |
|  | R | 5976 | 753 | 6322 | 879 | 6404 | 644 | 6785 | 590 | 6750 | 682 | 6727 | 553 |
| Thalamus (Female) | L | 5459 | 512 | 6033 | 1082 | 6510 | 836 | 6305 | 787 | 6547 | 713 | 6770 | 700 |
|  | R | 5370 | 516 | 5798 | 920 | 6323 | 815 | 6277 | 878 | 6267 | 561 | 6499 | 520 |
| Caudate | L | 3041 | 445 | 3180 | 466 | 3375 | 491 | 3479 | 533 | 3594 | 411 | 3616 | 487 |
|  | R | 2828 | 501 | 2795 | 595 | 2932 | 602 | 3106 | 657 | 3226 | 552 | 3272 | 548 |
| Putamen | L | 3953 | 554 | 4374 | 774 | 4613 | 746 | 5021 | 963 | 5344 | 618 | 5256 | 642 |
|  | R | 3904 | 720 | 4420 | 821 | 4634 | 763 | 4804 | 1087 | 5307 | 721 | 5239 | 709 |
| Pallidum | L | 1194 | 379 | 1241 | 417 | 1341 | 349 | 1542 | 400 | 1624 | 261 | 1642 | 280 |
|  | R | 1145 | 429 | 1134 | 470 | 1325 | 418 | 1393 | 459 | 1599 | 290 | 1622 | 303 |
| Hippocampus | L | 2686 | 312 | 2885 | 326 | 3149 | 322 | 3254 | 399 | 3427 | 312 | 3466 | 328 |
|  | R | 2594 | 268 | 2795 | 380 | 3008 | 273 | 3116 | 373 | 3308 | 263 | 3337 | 317 |
| Amygdala | L | 1036 | 194 | 1122 | 191 | 1193 | 217 | 1228 | 206 | 1251 | 222 | 1232 | 184 |
|  | R | 855 | 225 | 886 | 214 | 1005 | 192 | 1023 | 192 | 1123 | 179 | 1126 | 179 |
| Accumbens area | L | 484 | 109 | 630 | 183 | 631 | 174 | 697 | 168 | 651 | 120 | 653 | 171 |
|  | R | 577 | 208 | 780 | 266 | 836 | 268 | 855 | 244 | 776 | 192 | 774 | 221 |
| Ventral DC | L | 2715 | 420 | 3245 | 536 | 3410 | 561 | 3393 | 474 | 3268 | 490 | 3373 | 518 |
|  | R | 2659 | 399 | 3065 | 459 | 3293 | 505 | 3283 | 403 | 3199 | 440 | 3330 | 469 |

Note: The unit of volume is mL.

We examined the age-group difference in the volume of all subcortical regions. Figure 2B shows maps of mean annual change rates across 2-6, relative to the first year. The volume of subcortical regions increased with age. At age 6, the volume of putamen, palladium, amygdala, accumbens area, and cerebellum white matter was over 30% larger than age 1. The age-dependent volume change of subcortical regions was also symmetric. Figure 2D presents year-by-year changes (t-values) for all subcortical regions. According to these analyses, most subcortical regions’ volume significantly enlarged in age 2 and age 3. At age 4 and age 5, the volume of bilateral putamen, bilateral pallidum, and left amygdala further increased.

### Growth curve models and developmental patterns of brain morphometrics

We further modeled the age-dependent individual variability of brain morphometrics using the growth-curve model. Figure 3A presents examples of growth curve models for cortical thickness, subcortical volume, and brain tissue volume. The 5th, 10th, 25th, 50th, 75th, 90th, and 95th percentage curves were used to visualize the age-dependent distributions of individual variability. For a given chronological age and a morphometric feature, the growth curve model could derive a percentage value that indicates the relative position among the TDC distribution. Supplementary Figures 2-4 present growth curve models for the mean thickness of all cortical regions, the volume of all subcortical regions, and the volume of different types of brain tissues.

**Figure 3.**
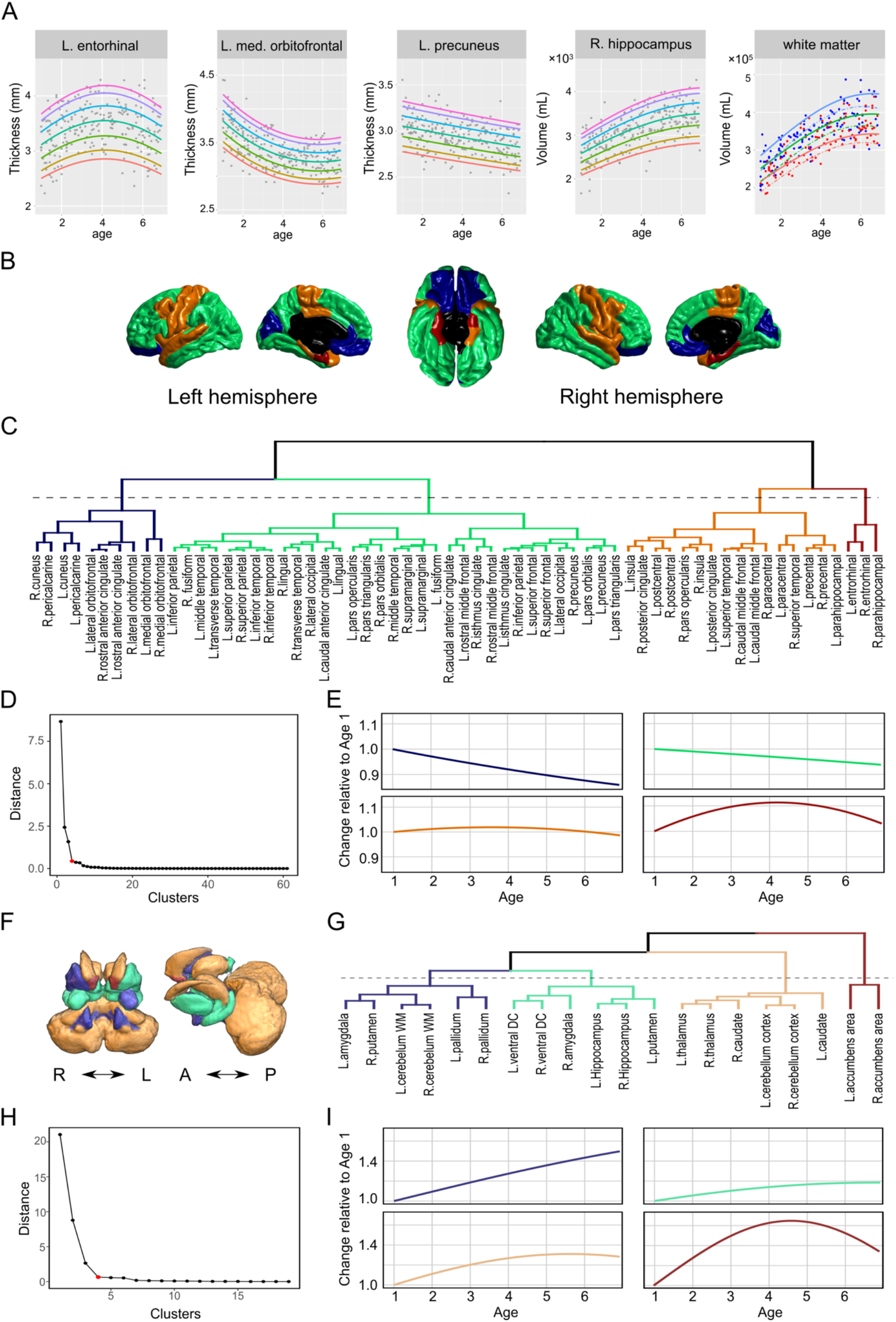
Regional growth curve models and clustering results of age-dependence patterns. A. Examples of regional growth curve models. The curves with different colors represent the 5^th^, 10^th^, 25^th^, 50^th^, 75^th^, 90^th^, and 95^th^ percentiles of the growth curves. The grey dots represent the actual data for fitting the models. For metrics showing significant sex differences, such as white matter volume, sex-specific growth curves are fitted. B. Visualization of the four types of cortical regions carrying different age-dependence patterns. C. Hierarchical dendrogram of similarity of growth curves among all cortical regions. D. A knee plot of inter-cluster distance as a function of the number of clusters in the dendrogram (panel C). Starting from 4 clusters (red dot), the distance is stable. E. Change rates of the median of the growth curves corresponding to the four clusters. F. Visualization of the four types of subcortical regions carrying different age-dependence patterns. G. Hierarchical dendrogram of similarity of growth curves among all subcortical regions. H. A knee plot of inter-cluster distance as a function of the number of clusters in the dendrogram (panel G). The distance is stable, starting from 4 clusters (red dot). I. Change rates of the median of the growth curves corresponding to the four clusters.

To further characterize the developmental patterns of regional brain morphometrics, we clustered the growth curves into different types based on their changing trends with age. Figure 3B maps four types of cortical regions carrying different age-dependence trends. Figure 3C further depicts a hierarchical dendrogram of similarity of growth curves among all cortical regions. According to the knee plot of inter-cluster distance (Figure 3D), we chose to cut the dendrogram to form 4 clusters. Figure 3E shows the age-dependent change rates of the median of the growth curves corresponding to the four clusters. The first two types of cortical regions exhibited monotonous decreasing trends with increasing age, and the difference between the two was the slope. The third type showed a nearly flat growth curve, and the last type demonstrated an inverted-U shape of the growth curve.

For the subcortical regions, we also clustered their volumes’ growth curves into four types (Figure 3F-G) according to the knee plot of inter-cluster distance (Figure 3H). Figure 3I shows the corresponding growth curves of the four types. The first type of subcortical regions, including the left amygdala, right putamen, bilateral cerebellum white matter, and bilateral pallidum, exhibited a steep increasing slope with age. The second type, including bilateral ventral DC, right amygdala, bilateral hippocampus, and left putamen, showed a power function-like trend that approximates to 1.2 (change rate relative to age 1). The last two types exhibited inverted-U shapes of the growth curve, while the trend was more apparent in the bilateral accumbens area (type 4). These results revealed the diversity of the age-dependent changes of cortical and subcortical regions from 1 to 6 years old.

### Recognizing abnormal development based on brain morphometrics

For a given brain region of an individual, the growth curve model can provide an individualized inference by deriving a relative position, as reflected by a percentage value, among the fitted distributions of age-dependent individual variability. We examined the utility of this feature to recognize children with delayed language and speech disorders (DLSD). The brain morphometrics of 20 DLSD and 20 age-matched TDC (not included in the samples used to build the growth curve models) were input to the growth curve models to obtain the corresponding percentage values, which were used to train a linear discrimination classifier. In Figures 4A, we present an exemplar growth curve representing the right lateral occipital gyrus’s cortical thickness. The TDC and DSLD participants are marked using black and red circles in the plots. While most TDC fell within the 5-95% range, a large portion of DSLD participants was outside this range.

**Figure 4.**
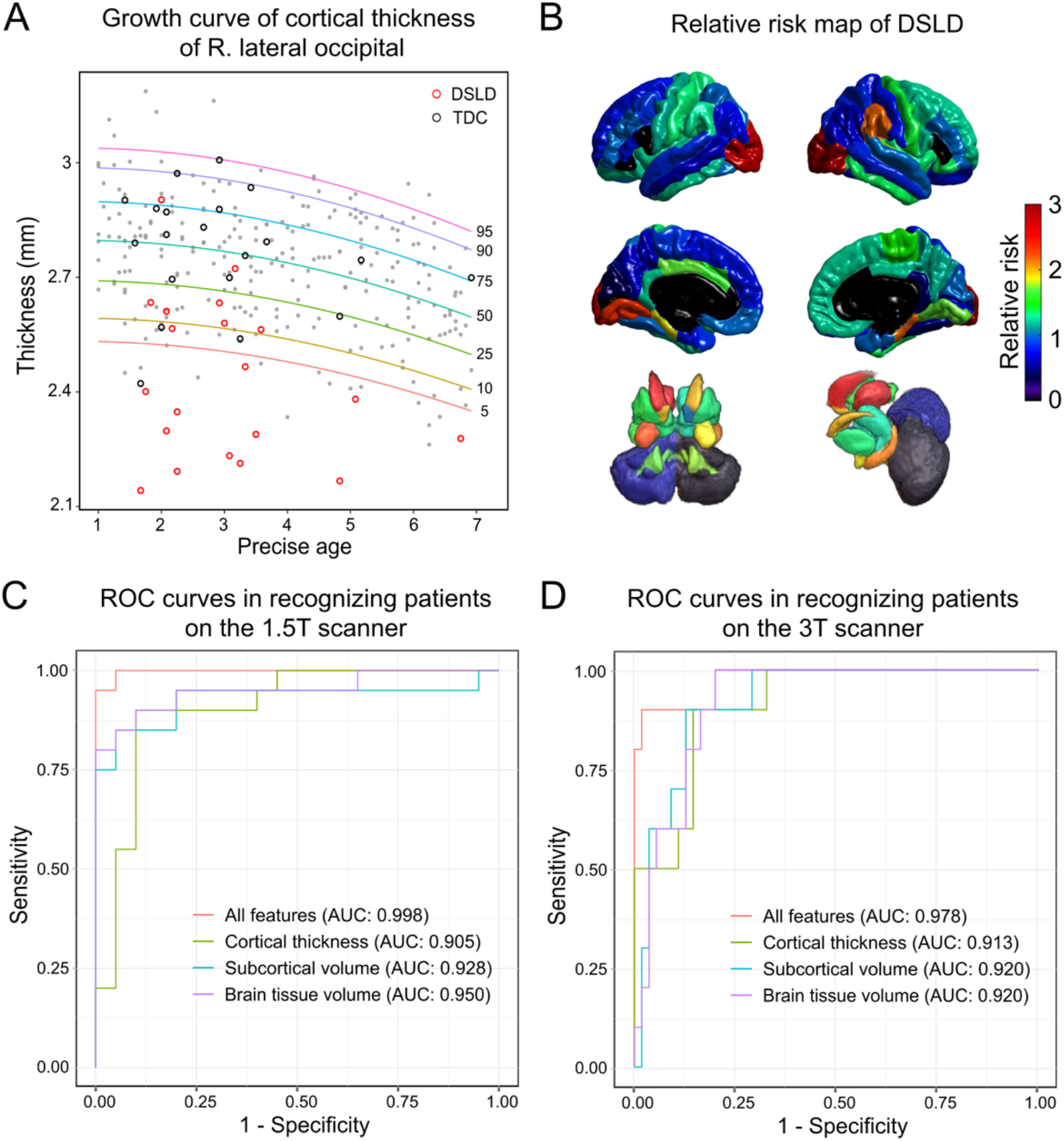
Multivariate classification of developmental speech and language disorder (DSLD) using growth curves of brain regions. A. A growth curve model for the cortical thickness of the right lateral occipital. The growth curve model is presented using curves indicating different percentiles in the population, from 5^th^ to 95^th^. The gray dots indicate data from individual participants that were used to construct the growth curve models. An independent set of 20 DSLD patients and 20 age-matched controls are presented using red and black circles, respectively. The locations of these circles indicate their position in the same-age population. B. Relative risk map of DSLD. The relative risk value for each region was calculated by dividing the proportion of DSLD among all participants showing abnormality by the proportion of DSLD among all participants not showing abnormality. The abnormality was defined as outside the 5%-95% range of the given region’s growth curve model. A high relative risk value indicates the abnormal development of a given brain region is a risk factor of DSLD. C. Receiver operation characteristic (ROC) curves of classifiers that identify DSLD patients from the typically developing children. These classifiers utilize the circles in the growth curve models (e.g., those in panel A) of brain morphometrics. The classifier that adopts the growth curve models of all morphological metrics, including regional thickness/volume metrics and brain tissue volume measures, achieves the highest area under the curve (AUC) of 0.998, with a classification accuracy of 0.975. D. ROC curves of the classifiers trained using the 1.5T data and tested using independent data from the 3T scanner. The AUC is 0.978 for the classifiers combining all the features, demonstrating the sensitivity and generalizability of the growth curves and DSLD recognition models.

To summarize the associations between DSLD and abnormal morphometrics detected in the regional growth curves, we present a relative risk map in Figure 4B. The value is a risk ratio of DSLD when a given region is abnormal (outside 5-95% range of the corresponding growth curve) versus when the region is normal. The thickness of the bilateral lateral occipital cortex, the right angular gyrus, the bilateral parahippocampal gyrus, the bilateral cingulate gyrus, and the right frontal cortex exhibited a high risk ratio. Most subcortical regions, especially the caudate, hippocampus, and amygdala, showed an increased DSLD risk ratio. These observations reveal that wide-spread brain regions are related to DSLD, supporting strong associations between DSLD and abnormal brain development as reflected in the growth curves.

To further combine the information from different regions to achieve accurate recognition of DSLD, we trained linear discrimination classifiers based on the percentile values derived from the growth curve models. With a 10-fold cross-validation scheme, the analyses achieved a classification accuracy of 0.975 in classifying DLSD and TDC participants using all regional cortical thickness, subcortical volume, and brain tissue volume features. The classification accuracy was 0.900 when using the cortical thickness measures, which was 0.875 when using the subcortical volume measures, which was 0.900 when using the brain tissue volume features. The performance of the classifiers was summarized using ROC plots in Figure 4C. The area under curve (AUC) metrics for the ROC analyses were 0.998, 0.905, 0.928, and 0.950 for all morphometrics, cortical thickness only, the subcortical volume only, and brain tissue volume only. These observations reflect that the growth curves help to derive individualized evaluation of brain development status with clinical potentials.

### Independent examination of sensitivity and generalizability of the model

We further examined the sensitivity and generalizability of the DSLD recognition model with data from an independent 3T scanner. From the growth curve models constructed using the data from the 1.5T scanner, we derived regional percentile values of 55 TDC and 10 DSLD patients scanned on the 3T scanner. We then applied the pre-trained linear discrimination classifiers to recognize DSLD patients based on the percentile values. As shown in Figure 4D, the classifiers achieved high performance as reflected by the AUC of ROC curves. When combining all regions’ percentile values, the classifier achieved an AUC of 0.978; when only using cortical thickness, subcortical volume, brain tissue volume, the AUCs of the classifiers were between 0.913 and 0.920. These results from an independent scanner with different magnetic strength support that the growth curve models and the DSLD classifiers based on the percentile values can provide individualized inferences sensitive to DSLD and generalizable across the scanner.

## Discussion

For the first time, the present study quantified annual changes of cortical thickness and subcortical volume of typically developing children between 1 and 6 years old. The thickness in most cortical regions decreased at age 2, age 3, and age 6. Simultaneously, the volume for the precentral gyrus, entorhinal cortex, and parahippocampal gyrus showed a different, inverted-U shape of dynamics. The volume of most subcortical regions significantly enlarged in age 2 and age 3, and the volume of putamen, pallidum, hippocampus, and amygdala kept increasing at age 4 and age 5. With the growth curve models for the age-dependence of individual variability, we were able to discriminate four age-dependent patterns among cortical regions and subcortical regions, providing a summary of brain morphometrics’ developmental patterns.

The regional growth curve models enabled identifying DSLD patients with high sensitivity and specificity. Our independent test further supported the generalizability of the growth curve models and the classifiers based on them. To our knowledge, these are the first brain regional growth charts for children ages 1 to 6 years, a critical time window for diagnosing many developmental disorders. The accuracy of our model, when all features were combined, reached 0.975 for recognizing DSLD. Since the human brain reaches 95% of adult size by age 6 [6], our current study addresses a period during which brain size develops dynamically and substantially and provides a link to existing studies of structural brain maturation beyond six years-of-age [27]. The growth curve model generated in the present study offers a potentially valuable tool to measure and define the developmental trajectory of individual brains and to provide clinically relevant information.

Previous studies revealed the potential importance of brain maturation curves in the diagnosis of early neurodevelopmental disorders [5,28]. For instance, early-onset schizophrenia patients were found to exhibit abnormal acceleration of gray matter loss during development [4,29]; autistic children showed distinct patterns of cortex growth and white-gray matter contrast [30,31], abnormal amygdala volume dynamics [32], and extreme male-like patterns of interhemispheric connectivity [33]; children with ADHD have been found to have reduced subcortical volume [34], delayed cortical maturation [35], and altered hemisphere asymmetry[36]; fragile X syndrome is associated with aberrant prefrontal cortex maturation[37], and volumetric abnormalities in subcortical regions [38]. Notably, most studies were based on adolescents imaged after disorders were diagnosed. Few papers have reported brain imaging samples during childhood [14,39]; future longitudinal studies are essential to improve the first diagnosis and initiate earlier treatments for developmental disorders.

Besides characterizing the age-dependence and variabilities of typical brain morphometry development between 1 and 6 years of age, our study represents an initial effort towards establishing growth charts of brain morphometry in children. The WHO published growth curve standards for body height and weight for children in 2006 [24], and the growth chart model has been used in clinical practice for many years, but there are no childhood growth charts for the typically developing brain. The models presented in this study represent an initial effort to construct brain growth charts for children, which would enhance the precise diagnosis of developmental brain disease and might even contribute to individualized education in the future. Although the sample size in this study is insufficient to construct a clinically useful growth chart, the present results serve as a starting point. The precision of the preliminary growth chart model can be improved by incorporating additional samples. For instance, based on the current models, one could implement a web-based service to evaluate brain abnormalities of images uploaded by users, providing a percentile among the same age population for every brain region. This web-based service would learn from the new data contributed and improve its precision.

One limitation of the present study is that the growth curve models were not validated using independent scanners. Future efforts will involve developing a common algorithm for different clinical scanners and fitting brain growth curve models to other racial/ethnic samples. The estimation procedure should consider scanner type and scanning parameters to provide clinically useful information across different centers. It would also be preferable to develop an automatic detection algorithm for other neurological or psychiatric disorders during development, which would require longitudinal data collection with larger sample size, including sufficient subgroups of different types of patients.

In summary, this study described easy-to-use structural MRI-based brain regional growth charts, with the potential to predict brain developmental stage and clinical aberrance.

## Data Availability

The imaging data used in this study are available upon direct request to the corresponding authors. The request should be reviewed and approved by the institutional ethics committee of Shanghai Mental Health Center and Children's Hospital, Zhejiang University.
The code for analyzing the data is available at https://github.com/yangzhi-psy/braindev1_6, and is free to download and re-use with proper citations.
The MRI brain templates for children from 1 to 6 years old (3D nifty files) are shared in a public open-science repository.

https://osf.io/fm7cq/?view_only=9716e89f09e04b4bb2b4f0323ab2b684.

## Acknowledgements

The authors acknowledge Dr. Xiaolu Ruan and Ms. Jingjing Liu for their help in cleaning up the data.

## Funds

This work was supported by the National Key Research and Development Program of China (2018YFC2001600, 2016YFC1305301, 2016YFC1306205); National Natural Science Foundation of China (81971682, 81571756, 81270023, 81570759, 81270938, 81573516); Natural Science Foundation of Shanghai (20ZR1472800); Shanghai Municipal Commission of Education-Gaofeng Clinical Medicine Grant Support (20171929); Hundred-Talent Fund from Shanghai Municipal Commission of Health (2018BR17); Scientific Research Fund of Zhejiang Provincial Education Department (Y201431325); Shanghai Mental Health Center Clinical Research Center (CRC2018DSJ01-5; CRC2019ZD04); Research Funds from Shanghai Mental Health Center (13dz2260500, 2018-YJ-02).

