## Supplemental for "Growth charts for individualized evaluation of brain morphometry for preschool children"

### Supplementary Materials

**Supplementary Figure 1.** A flowchart of the image processing pipeline. The pipeline consists of 5 steps, as represented by different color blocks, including template construction, image segmentation, surface reconstruction, modeling, and performance test. The black arrows depict the procedures to construct the normative growth curve model. The red arrows indicate the processes to evaluate the performance of the model to recognize developmental disorder individuals. To increase the accuracy of the "Individual WM/GM/CSF segmentation", we build age-specific brain templates using ten male and ten female subjects in each age group and construct age-specific a priori images for segmentation. The customized individual tissue segmentation is then inserted into the Freesurfer pipeline to estimate cortical thickness and subcortical volumes. Based on these features, we construct regional growth curve models and train brain-age prediction models. The performance of these models to reveal brain development deficits is examined using an independent dataset.

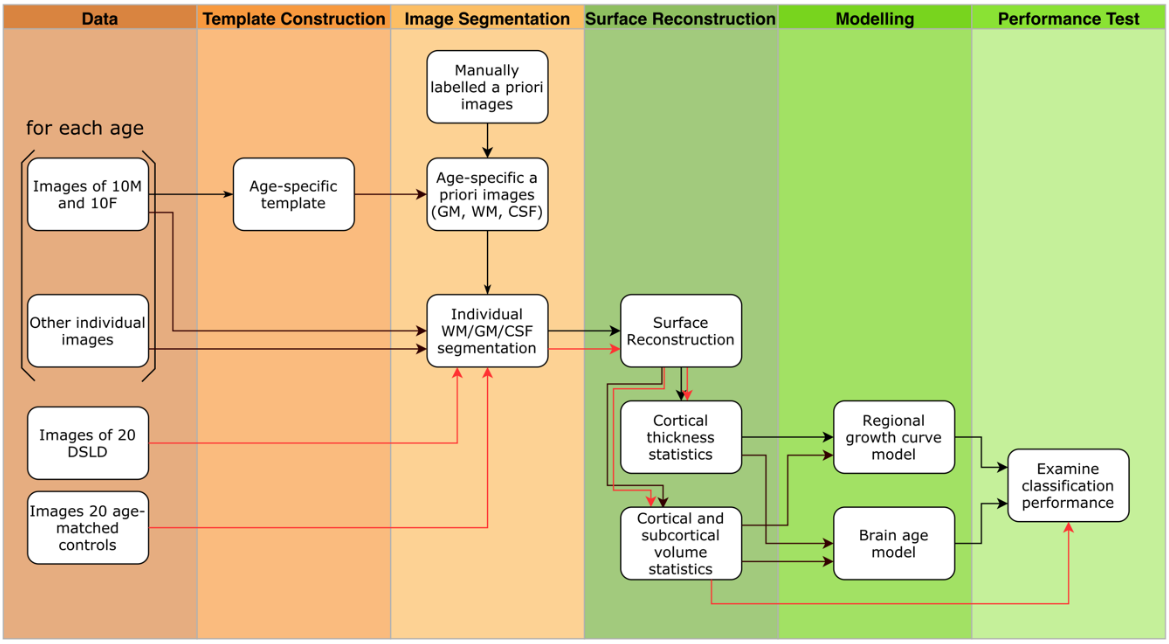

**Supplementary Figure 2.** Growth curve models of left hemisphere cortical regions

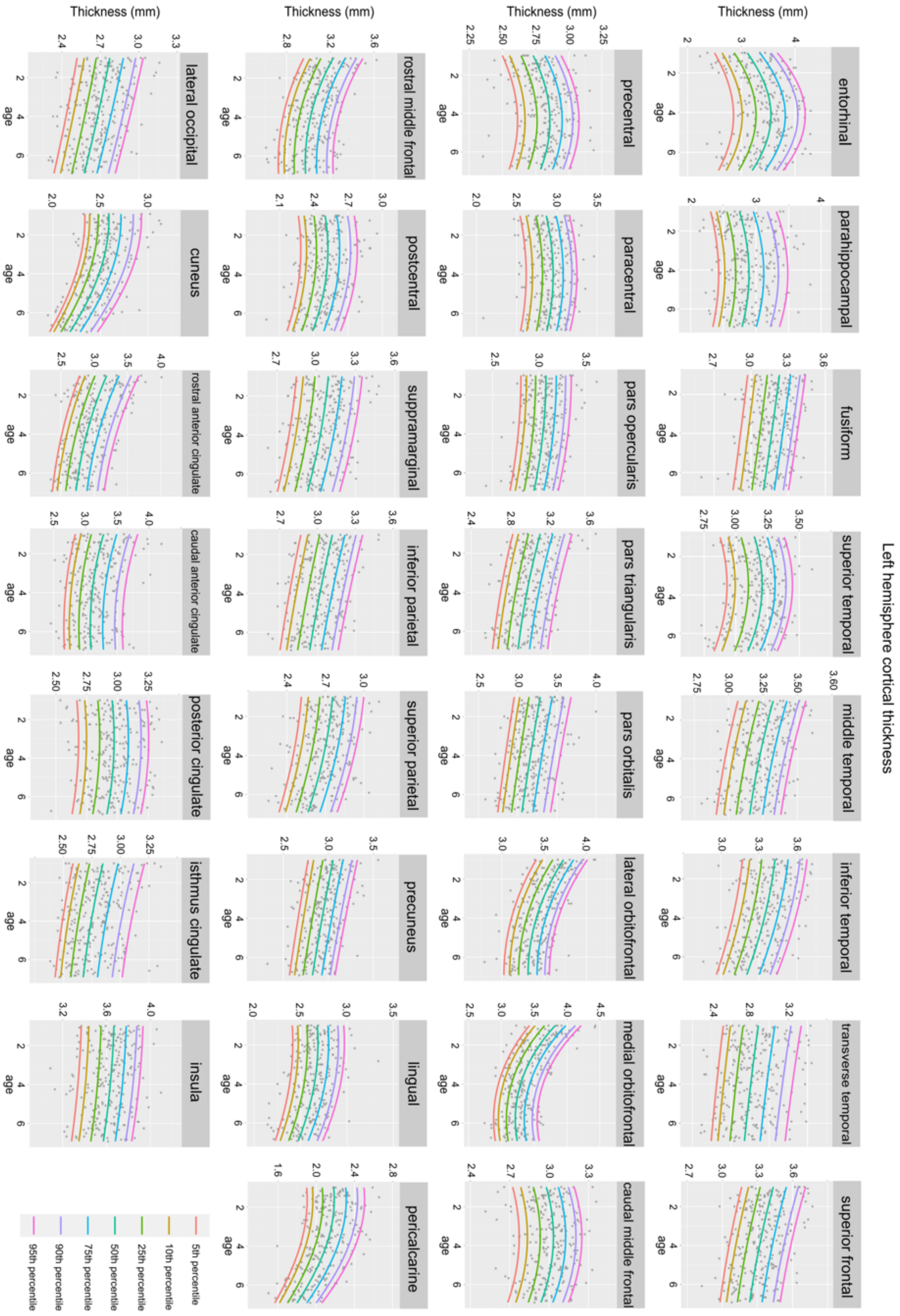

**Supplementary Figure 3.** Growth curve models of right hemisphere cortical regions

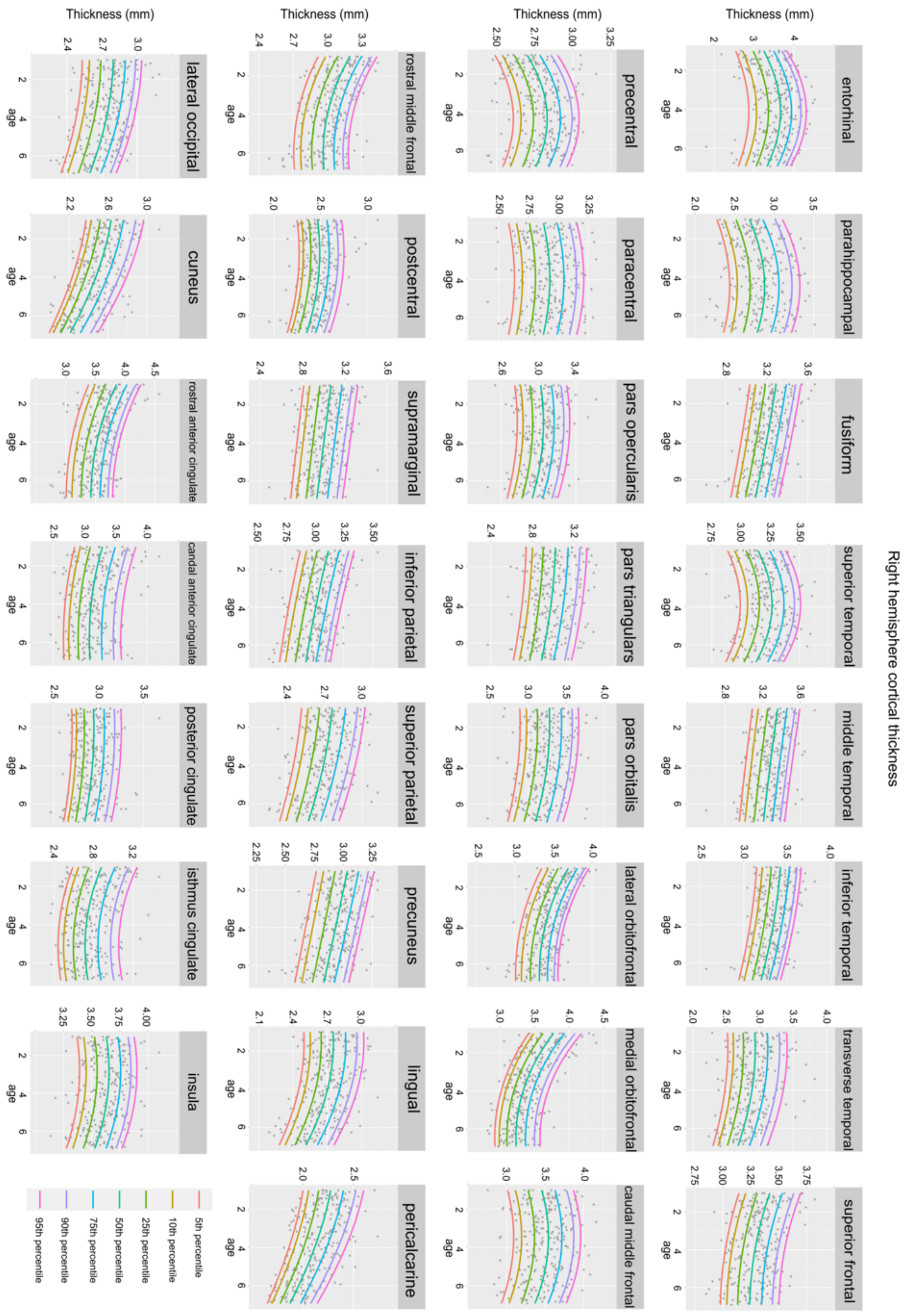

**Supplementary Figure 4.** Growth curve models of subcortical regions and brain tissues

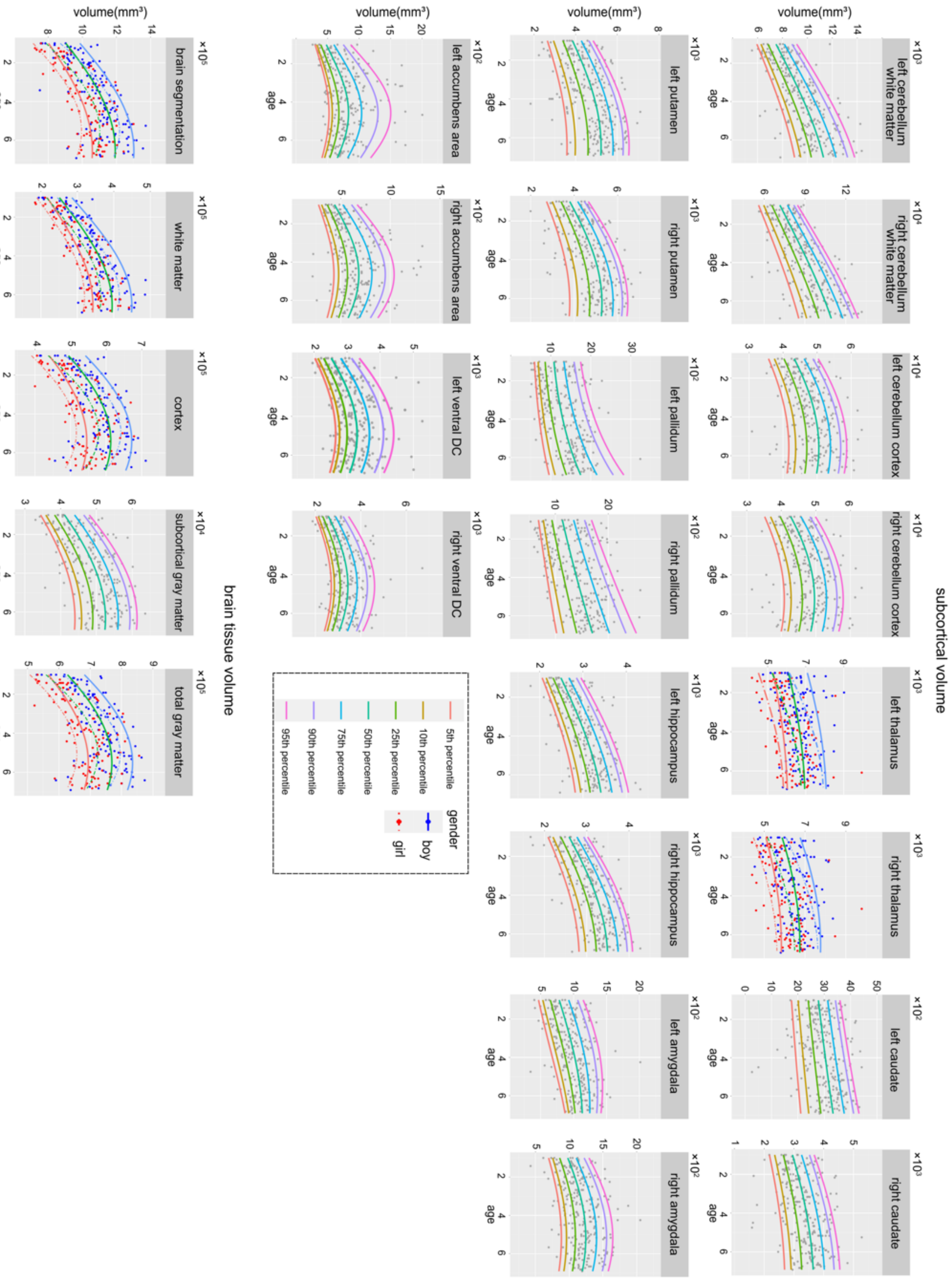

**Supplementary Table 1.** Complaints of typically developing children scanned in 1.5T MRI

| Complaints | Number of Males | Age of Male | Number of Females | Age of Female | Total Number |
|---|---|---|---|---|---|
| One episode of seizure | 53 | 1-6.25 | 40 | 1-6.75 | 93 |
| Headache | 40 | 1.92-6.92 | 44 | 1.33-6.83 | 84 |
| Physical examination | 21 | 1.5-5.17 | 15 | 1.25-6.25 | 36 |
| Dizziness | 16 | 1-6.58 | 11 | 1.42-6.58 | 27 |
| Facial paralysis | 9 | 1-6.75 | 6 | 1.75-5.08 | 15 |
| Trauma | 5 | 2.5-5.75 | 0 | - | 5 |
| Short-term fever of unknown | 3 | 1-2 | 2 | 2.41-3.25 | 5 |
| Total | 135 | 1-6.92 | 110 | 1-6.83 | 265 |

**Supplementary Table 2.** Complaints of typically developing children scanned in 3T MRI

| Complaints | Number of Males | Age of Male | Number of Females | Age of Female | Total Number |
|---|---|---|---|---|---|
| One episode of seizure | 11 | 1.04-3.75 | 17 | 1.13-5 | 28 |
| Headache | 7 | 3.04-6.87 | 5 | 5.45-6.78 | 12 |
| Physical examination | 2 | 1.17-3.82 | 6 | 1.17-6.80 | 8 |
| Dizziness | 1 | 6.09-6.09 | 2 | 4.72-6.31 | 3 |
| Facial paralysis | 1 | 1.87-1.87 | 0 | - | 1 |
| Short-term fever of unknown | 0 | - | 3 | 1.48-2.85 | 3 |
| Total | 22 | 1.04-6.87 | 33 | 1.13-6.80 | 55 |

**Supplementary Table 3.** Age-dependence of brain tissues, stratified by sex

| Age group | Statistics | Brain volume, excluding ventricles | | Total gray matter volume | | Total cortex volume | | Total cerebral white matter volume | |
|---|---|---|---|---|---|---|---|---|---|
| | | M | F | M | F | M | F | M | F |
| Age1 | Mean[a] | 921006 | 846052 | 641623 | 595331 | 507171 | 468799 | 263700 | 236739 |
| | SD[a] | 90264 | 64983 | 56498 | 40755 | 48412 | 36002 | 37271 | 24998 |
| Age2 | Mean[a] | 1030867 | 927474 | 702509 | 638696 | 559802 | 504513 | 312542 | 273324 |
| | SD[a] | 75588 | 76178 | 47895 | 51885 | 47275 | 45242 | 32490 | 25441 |
| | Change rate[b] | 1.10 | 1.12 | 1.07 | 1.09 | 1.08 | 1.10 | 1.15 | 1.19 |
| | t-val[c] | 5.08*** | 3.71*** | 4.47*** | 2.99*** | 4.22*** | 2.81*** | 5.37*** | 4.69*** |
| Age3 | Mean[a] | 1079276 | 1038915 | 730148 | 698590 | 580949 | 551940 | 340729 | 322632 |
| | SD[a] | 93707 | 83692 | 43914 | 51549 | 38224 | 48197 | 29431 | 34743 |
| | Change rate[b] | 1.23 | 1.17 | 1.17 | 1.14 | 1.18 | 1.15 | 1.36 | 1.29 |
| | t-val[c] | 2.14* | 4.34*** | 2.27* | 3.62*** | 1.86 | 3.16*** | 3.44*** | 5.04*** |
| Age4 | Mean[a] | 1153792 | 1073595 | 764143 | 718080 | 608458 | 575960 | 371279 | 339398 |
| | SD[a] | 45589 | 85528 | 31608 | 54921 | 25363 | 52990 | 21394 | 34754 |
| | Change rate[b] | 1.27 | 1.25 | 1.21 | 1.19 | 1.23 | 1.20 | 1.43 | 1.41 |
| | t-val[c] | 3.62*** | 1.19 | 3.08*** | 1.06 | 2.97*** | 1.37 | 4.12*** | 1.40 |
| Age5 | Mean[a] | 1163194 | 1101837 | 758841 | 725251 | 601626 | 580387 | 385438 | 359403 |
| | SD[a] | 100797 | 79164 | 60489 | 50705 | 56553 | 45774 | 43639 | 33780 |
| | Change rate[b] | 1.30 | 1.26 | 1.22 | 1.18 | 1.24 | 1.19 | 1.52 | 1.46 |
| | t-val[c] | 0.41 | 0.98 | -0.38 | 0.39 | -0.54 | 0.25 | 1.41 | 1.67 |
| Age6 | Mean[a] | 1170756 | 1095470 | 756045 | 707478 | 601031 | 557665 | 396089 | 367898 |
| | SD[a] | 134124 | 78152 | 82877 | 51954 | 75113 | 44250 | 56588 | 33630 |
| | Change rate[b] | 1.29 | 1.27 | 1.19 | 1.18 | 1.19 | 1.19 | 1.55 | 1.50 |
| | t-val[c] | 0.19 | -0.26 | -0.12 | -1.11 | -0.03 | -1.61 | 0.64 | 0.81 |

Note:

a. volume unit: mL;

b. change rate relative to age 1;

c. t values relative to the previous age-group; *** false discovery rate $q<0.05$;** $p<0.01$; * $p<0.05$.
